# Selective Posterior Cerebral Artery Wada Better Predicts Good Memory and Naming Outcomes Following Selective Stereotactic Thermal Ablation for Medial Temporal Lobe Epilepsy Than Internal Carotid Artery Wada

**DOI:** 10.1101/2024.03.24.24304488

**Authors:** Daniel L. Drane, Emma Acerbo, Anna Rogers, Nigel P. Pedersen, Adam Williamson, Matthew A. Stern, Adam S. Dickey, Brian M. Howard, Donald J. Bearden, Noah Okada, Ekaterina Staikova, Claire Anne Gutekunst, Abdulrahman Alwaki, Timothy Gershon, Viktor Jirsa, Robert E. Gross, David W. Loring, Ammar Kheder, Jon T. Willie

## Abstract

The conventional intracarotid amobarbital (Wada) test has been used to assess memory function in patients being considered for temporal lobe epilepsy (TLE) surgery. Minimally invasive approaches that target the medial temporal lobe (MTL) and spare neocortex are increasingly used, but a knowledge gap remains in how to assess memory and language risk from these procedures. We retrospectively compared results of two versions of the Wada test, the intracarotid artery (ICA-Wada) and posterior cerebral artery (PCA-Wada) approaches, with respect to predicting subsequent memory and language outcomes, particularly after stereotactic laser amygdalohippocampotomy (SLAH). We included all patients being considered for SLAH who underwent both ICA-Wada and PCA-Wada at a single institution. Memory and confrontation naming assessments were conducted using standardized neuropsychological tests to assess pre- to post-surgical changes in cognitive performance. Of 13 patients who initially failed the ICA-Wada, only one patient subsequently failed the PCA-Wada (p=0.003, two-sided binomial test with *p*_0_=0.5) demonstrating that these tests assess different brain regions or networks. PCA-Wada had a high negative predictive value for the safety of SLAH, compared to ICA-Wada, as none of the patients who underwent SLAH after passing the PCA-Wada experienced catastrophic memory decline (0 of 9 subjects, *p*<.004, two-sided binomial test with *p*_0_=0.5), and all experienced a good cognitive outcome. In contrast, the single patient who received a left anterior temporal lobectomy after failed ICA- and passed PCA-Wada experienced a persistent, near catastrophic memory decline. On confrontation naming, few patients exhibited disturbance during the PCA-Wada. Following surgery, SLAH patients showed no naming decline, while open resection patients, whose surgeries all included ipsilateral temporal lobe neocortex, experienced significant naming difficulties (Fisher’s exact test, *p*<.05). These findings demonstrate that (1) failing the ICA-Wada falsely predicts memory decline following SLAH, (2) PCA-Wada better predicts good memory outcomes of SLAH for MTLE, and (3) the MTL brain structures affected by both PCA-Wada and SLAH are not directly involved in language processing.

## Introduction

Recent years have witnessed the proliferation of innovations in the surgical management of epilepsy such as robot-assisted stereoelectroencephalography, stereotactic laser interstitial thermal therapy, radiofrequency [RF] ablation, neuromodulation, and focused ultrasound.^1-5^ Highly focal ablations and network targeting therapies require more precise localizations of epileptogenic zones as well better assessments of associated cognitive risks of surgery. A traditional technique for determining the safety of epilepsy surgery in patients with temporal lobe epilepsy (TLE) has been the intracarotid artery amobarbital (ICA-Wada) procedure, which predominantly targets the lateral frontal lobe and anterior-lateral temporal lobe.^6^ Although fMRI has generally supplanted the ICA-Wada test for assessing language dominance,^7^ the test is still often used to assess the capability of unanesthetized brain (particularly the hemisphere contralateral to the seizure onset zone) to sustain memory function and to predict and avoid catastrophic memory outcomes after surgery for TLE. In this context, a “failed” Wada refers to inability of the unanesthetized brain to support memory. By contrast, the selective posterior cerebral artery Wada (PCA-Wada) targets the medial temporal lobe including hippocampus, but has not been widely practiced due to a need for greater technical expertise and a perceived elevated procedural risk of stroke. A question that arises is whether the ICA- or PCA-Wada accurately predict memory and naming deficits associated with certain temporal lobe surgeries. In cases of “failed” ICA-Wada it has the practice of our institution to then perform PCA-Wada to further assess risk. Here we report the neurocognitive outcomes of all patients undergoing temporal lobe surgery after first undergoing both ICA- and PCA-Wada tests.^6,8,9^ We also discuss emergent approaches with electrical stimulation that may eventually replace pharmacological approaches.

In the conventional Wada test, a barbiturate (e.g., sodium amobarbital, methohexital) is injected into the ICA to anesthetize portions of the ipsilateral hemisphere to assess memory function of the contralateral hemisphere, which is generally unaffected. However, several studies have demonstrated that brain regions affected by barbiturate injection can vary due to inconsistent ICA hemodynamics diluting the dose delivered to the hippocampus. Additionally, ICA Wada may not anesthetize the entire hippocampus, which has variable blood supply.^10-14^ The arterial supply to the hippocampus is comprised of branches of the anterior choroidal artery (AchorA), which derives from the ICA, and one or more hippocampal arteries from the PCA that make an anastomotic arcade along the anteroposterior axis of the hippocampus.^15^ Figure 1 schematically represents this anatomy. Indeed, Urbach et al.^11,12^ have demonstrated that the ICA Wada test lacks distribution to the posterior two-thirds of the hippocampus and de Silva et al.^13^ showed that the regions of the temporal lobe affected by the ICA Wada are not limited to the hippocampus.

**Figure 1.**
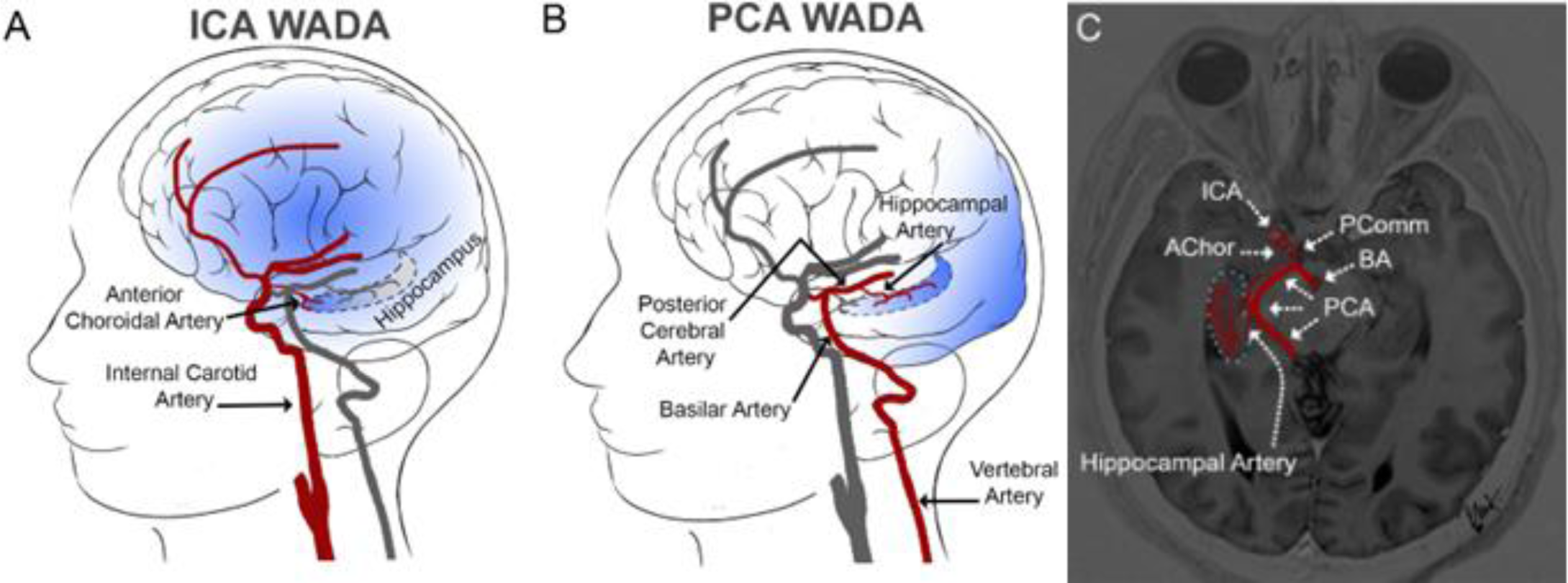
ICA vs. PCA Vascular Territories in the Wada Test. **A.** This schematic depicts the relevant vascular anatomy of the ICA and PCA injections sites during the Wada test and highlights the expected regional differences in anesthetizatized territories. **A**. Infusion of barbiturate into the ICA temporarily reduces neuronal activity broadly across the cerebral hemisphere, including the anterior and middle cerebral artery distributions. The blood supply to the medial temporal lobe and hippocampus is variable with contributions from branches of the anterior choroidal artery from the ICA. **B**. The PCA supplies blood to portions of the medial temporal lobe, including the hippocampus via the hippocampal artery or arteries, which may derive from the cisternal segment of the PCA directly, or as a single or multiple branches of any of the inferior temporal arteries, which include the anterior, middle and posterior temporal arteries. **C**. An oblique slice through the hippocampus is outlined by light blue dots with blood supply via both the ICA and PCA distributions depicted in the axial view of an MPRAGE sequence MRI. (See supplemental materials for additional illustrative angiogram images and discussion).

The hippocampus is a potential target of the Wada test because it has been linked to memory encoding and retrieval,^16^ is often a source of ictal activity,^17^ and is a common target among surgical candidates.^18^ Nevertheless, a thorough consideration of extant animal and human research demonstrates that memory is supported by structures in addition to the hippocampus,^19-22^ and surgery involving a range of extra-hippocampal regions, despite sparing of the hippocampus, can result in memory decline.^23-25^ In particular, we have reported significant declines on standard verbal memory tasks following focal thermal ablations restricted to TL neocortices that spare medial TL regions. Although clinical decision making has historically emphasized the role of hippocampus in memory, the neocortices of the TL and frontal lobe must contribute as well.

The ICA-Wada test was developed following the well-known clinical case study of patient H.M., who became amnestic after undergoing bilateral medial TL resections for epilepsy.^6,26,27^ This case launched modern memory theories, which have focused largely on the contribution of the hippocampus to memory function. Juhn Wada developed the test that bears his name to predict and avoid amnestic outcomes.^6^ The Wada test predicts postoperative deficits by formally assessing the integrity of the contralateral (i.e. unoperated) temporal lobe to support memory function in the simulated absence of ipsilateral (i.e. operated) anterior temporal lobe function. As the open ATL was the primary procedure employed for decades, the ICA-Wada was a reasonable approach to assessing risk of deficits after surgery. Over time, however, the ICA-Wada has been criticized as a potentially unreliable predictor of postsurgical memory outcome. Despite these limitations,^28^ many clinicians still trust its utility in avoiding catastrophic memory decline, especially given the rise of alternative surgical strategies and neuromodulation.

It has long been the practice of our center to use the PCA-Wada whenever an ICA-Wada failed on technical grounds (e.g., angiographic cross-filling of contrast to the contralateral TL due to atypical vascular anatomy). Additionally, in the setting of surgery targeting only the MTL, we also considered the possibility that the ICA-Wada might produce false positive prediction of memory deficits due to mismatch between the vascular territories being interrogated and the epilepsy zones to be surgically ablated. Especially after initiating use of the stereotactic laser amygdalohippocampotomy (SLAH) approach,^4^ a procedure that targets the MTL while sparing anterior and lateral temporal structures (Figure 2), we began routinely obtaining the PCA-Wada whenever the ICA-Wada indicated memory failure, believing the PCA-Wada to better interrogate the medial TL to be surgically targeted. The PCA Wada preferentially delivers barbiturate to the MTL via the P2 segment of the PCA, with likely less off-target suppression of the ipsilateral TL neocortex and frontal lobe. Indeed, the PCA supplies blood to the medial and posterior portions of the hippocampus and single photon emission computed tomography (SPECT) studies have shown radiotracer distribution involving the entire hippocampus, parahippocampal gyrus, and occipital lobe.^11^ Historically, the PCA-Wada has been regarded as having a higher risk of procedural complications than the ICA-Wada, as PCA injection requires positioning the catheter deeper into the cerebral circulation; however, modern data suggests that the risks are quite low in experienced centers with appropriate patient selection (e.g., avoidance in patients with elevated stroke risk).^8^ Another recent study likewise relied upon selective PCA-Wada to evaluate risk prior to open subtemporal selective amygdalohippocampectomy (SAH).^29^ This group successfully used the PCA-Wada in 33 patients who failed the ICA-Wada, and 13 went on to undergo subtemporal SAH without significant adverse effects on memory.

**Figure 2.**
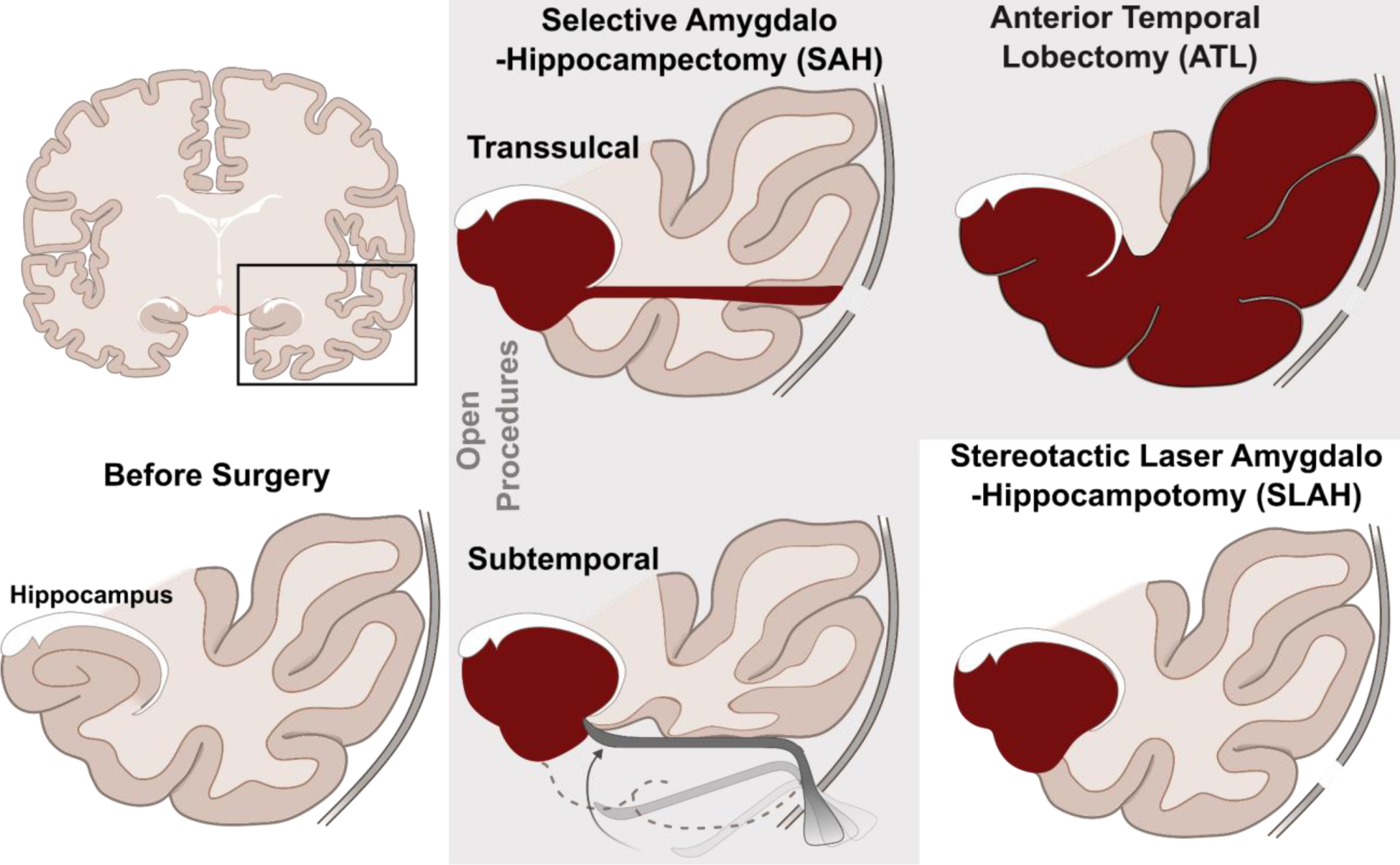
Illustration of distinct surgical approaches to medial temporal lobe epilepsy. Open selective amygdalohippocampectomy (SAH) is a surgical procedure that specifically targets and removes medial TL (particularly the amygdala, hippocampus, and parahippocampus/entorhinal cortex) but either transects (via trans- or para-sulcal approaches) or acutely retracts (subtemporal approach) to gain medial access. The subtemporal approach has been purported to better preserve the white matter in the temporal lobe. By comparison, open anterior temporal lobectomy (ATL) is a surgical procedure that removes the medial TL by first resecting the anterior-lateral TL and the temporal pole. Finally, stereotactic laser amygdalohippocampotomy (SLAH) is a minimally invasive approach that uses a small optical fiber to perform laser thermal interstitial therapy to precisely ablate the medial TL with negligible disturbance to anterior, basal, or lateral TL structures. Open procedures are grouped in a gray box.

In the present study, we present a series of TLE patients who underwent both ICA- and PCA-Wada tests as part of their work-up for epilepsy surgery. Overall, we hypothesized that:^1^

1. Only a minority of patients who failed the ICA-Wada would subsequently fail the PCA- Wada because the ICA-Wada would more broadly impact memory-involved structures beyond the MTL.
2. No patients who underwent SLAH after a failed ICA-but passed PCA-Wada would exhibit a catastrophic memory outcome. That is, the ICA-Wada is hypothesized to overestimate risk of memory decline with SLAH. In contrast, the PCA-Wada should better predict memory outcome after SLAH than the ICA-Wada due to closer concordance between the anesthetized brain region and the subsequent ablation.
3. Language disturbance would not be seen in SLAH patients either during the PCA-Wada nor during post-operative assessment, but would be more likely to occur in open resection procedures that extend beyond the MTL and involving neocortex supporting broader language network (e.g., anterior-basal and anterior-lateral temporal lobe – regions that would typically be included in a standard anterior temporal lobectomy [ATL] procedure) (Figure 2).

As the PCA-Wada has not been extensively studied, we provide detailed clinical findings in patients who underwent this test following a failed ICA-Wada. We sought to validate the utility of both the ICA- and PCA-Wada tests by examining procedure complications and memory outcomes in TLE patients who underwent subsequent epilepsy surgery, especially SLAH. We successfully treated patients who failed the conventional ICA-Wada but subsequently passed the PCA-Wada. These data also inform our team’s ongoing efforts to use direct electrical stimulation as an “electric Wada.”^30,31^

## Methods

### Participants

With approval from the Emory University Institutional Review Board, we retrospectively reviewed all patients in the Emory University Epilepsy Center who had ICA-Wada administered over a seven-year span that we had available. For this study, we included the subset of patients who were also administered a PCA-Wada after ‘failing’ the ICA Wada (failure criteria described below). For included patients, we reviewed demographic and neuropsychological testing reports that were performed at the following time points: pre-surgical baseline, then 6-8 and 12-18 months after surgery. The following memory tests were evaluated for significant change: Rey Auditory Verbal Learning Test,^32^ Visual Reproduction and Logical Memory subtests of the 4^th^ edition of the Wechsler Memory Scale,^33^ and the Rey Complex Figure Test.^34^ All patients also completed the Boston Naming Test (BNT).^35^

### Catheterization and injection

Following local anesthesia, arterial access was gained by Seldinger technique. The ICA was catheterized with a 5 French diagnostic catheter over a guidewire. A series of diagnostic cerebral angiograms were then completed. Injections were performed at the typical rate (8-10cc/second) to evaluate the intracranial vasculature including patency of the anterior and posterior communicating arteries. Cross filling to the contralateral hemisphere was evaluated on the initial injection. A second injection was then completed at the rate of anticipated drug infusion (2-4cc/second) to assure that cross filling to the contralateral hemisphere did not occur, which would confound the Wada test by anesthetizing parts of both hemispheres. Sodium amytal (100-125 mg), was injected intra-arterially into the ICA. Motor examination and electroencephalogaphy (EEG) were performed to confirm that the target hemisphere was anesthetized. Neuropsychological examination was then performed.

Following a ‘failed’ ICA Wada test, and after an adequate time for drug washout and neurological recovery, a PCA Wada test was completed during the same session. The targeted vertebral artery was catheterized using a diagnostic catheter and guidewire in a fashion similar to the ICA. A posterior circulation cerebral angiogram was completed. The diagnostic catheter was then exchanged for a guide catheter through which a 0.017” microcatheter was coaxially inserted into the PCA of interest over a 0.014” microguidewire. A PCA selective angiogram was then completed to assure that the microcatheter was distal to the brainstem and thalamic perforators. Additionally, the selective angiogram confirmed opacification of the inferior temporal branches of the P2 segment, including the hippocampal, anterior, middle and posterior temporal arteries. Once the appropriate anatomy was affirmed, 50-75 mg of sodium amytal was injected through the microcatheter to temporarily anesthetize the structures of the medial temporal lobe.

### Wada memory protocol

We used the Medical College of Georgia (MCG) Wada procedure for all patients.^36^ This procedure involves exposure to eight novel, three-dimensional objects while under the effects of the barbiturate, followed by a recognition recall of the eight objects presented in a set of foils (24 total objects) after resolution of drug effects. Immediately following barbiturate injection, eight 3-dimensional objects were shown to the patient, with each presented across both visual fields to the patient for approximately 5 seconds. After the patient returned to baseline as evidenced by a restoration of motor and language function and normalization of scalp EEG, the patient was shown the same eight objects, in a mixed order with 16 novel objects, and asked to verbally confirm if they had seen each object. Ipsilateral failure was defined as < 3/8 correctly recognized objects after a correction factor was applied for false positive errors. In the case of a failed ICA Wada, the patient underwent subsequent PCA Wada in the target hemisphere and the same protocol for neuropsychological testing was performed with a different set of objects.

### Statistical Approach

We provide detailed demographic and disease-related information at the individual level for each patient in our series. For hypotheses 1 and 2, we used a binomial test to compare the proportion of patients with a failed PCA Wada (#1) or catastrophic memory outcome (#2) to an assumed baseline event rate of 50%. For hypothesis 3, we used a Fisher’s exact test to compare the rate of decline in naming ability in patients who underwent a SLAH versus an open resection. We also compared pre-to post-surgical memory performances to determine if there were any significant changes in performance following surgery at the individual level. We used reliable change indices for a non-normally distributed test (i.e., the BNT) and otherwise employed a standard deviation method for assessing significant change. Finally, we computed a percent change in the raw scores for neuropsychological tests pre- and post-surgery and used an unpaired t-test to compare the % change for SLAH versus open surgery.

## Results

Overall, out of 82 patients who had an ICA-Wada during the six-year time span examined, 13 patients (15.9%) failed and went on to have a PCA Wada (Table 1). Details of the PCA-Wada are presented in Table 2. Of the PCA-Wada patients (2 male), all but two were left language dominant, 9 had ipsilateral mesial temporal sclerosis, and approximately 5 of 13 had poor baseline ipsilateral memory scores (i.e., > 2 SD below normal). Typically, the multidisciplinary epilepsy conference consensus was to obtain the ICA-Wada when other discordant factors pointed to possible compromise of the contralateral–presumably less involved–hemisphere (e.g., neuropsychological profile lateralized more to the contralateral side or appeared bilaterally decreased; bilateral structural MRI or PET hypometabolic abnormalities). No procedural complications occurred with the PCA-Wada at our center. Expected transient clinical changes during the PCA-Wada included hemianopsia in most subjects (8/12; 75%), but rarely included hemiplegia (1/13; 7.7%) or language disturbance (2/13; 15.4%). As noted, 12 of 13 patients who failed the ICA-Wada passed the PCA-Wada. Concurrent EEG confirmed adequate anesthetization of target brain regions for both the ICA and PCA.

**Table 1.** Demographics of all surgical patients who underwent a PCA Wada after failing the ICA Wada.

| Patient | Sex | Range Age | Range Age of SZ Onset | SZ Type | Surgery Type | MRI/PATH | Handedness | Language Dominance |
| --- | --- | --- | --- | --- | --- | --- | --- | --- |
| 1 | F | 50-55 | 30-35 | FIAS, secondary GTC | 1) Right SLAH<br>2) Right SAH | Normal | Left | Bilateral (Left>Right) |
| 2 | F | 50-55 | 10-15 | FIAS | Left SLAH | Left MTS | Right | Left |
| 3 | F | 40-45 | 35-40 | FIAS | Left SLAH | Left MTS | Right | Left |
| 4 | F | 45-50 | 20-25 | FIAS | Left SLAH | Left MTS | Right | Left |
| 5 | F | 18-20 | 15-20 | FIAS | Left SLAH | Normal | Right | Left |
| 6 | M | 50-55 | 25-30 | FIAS | Left SLAH | Left MTS | Right | Left |
| 7 | M | 40-45 | 15-20 | FIAS, rare GTCs | Left SLAH | Left MTS | Right | Left |
| 8 | F | 30-55 | 25-30 | FIAS | Left SLAH | Normal | Right | Left |
| 9 | F | 40-45 | 5-10 | FIAS, GTC, Myoclonic | Right SLAH | Right MTS | Right | Bilateral (Left>Right) |
| 10 | F | 55-60 | 15-20 | FIAS | Left SAH | Left MTS | Right | Left |
| 11 | F | 60-65 | 55-60 | FIAS | Left ATL | Normal | Left | Left |
| 12 | F | 40-45 | 15-20 | FIAS | Left RNS | Normal | Right | Left |
| 13 | F | 45-50 | 10-15 | FIAS, secondary GTC | Right RNS | Normal | Left | Right |

Note. PCA = posterior cerebral artery; ICA = internal carotid artery; SZ = seizure; M = male; F = female; FIAS = focal impaired awareness seizure; GTC = generalized tonic-clonic seizure; SLAH = stereotactic laser amygdalohippocampotomy; SAH = selective amygdalohippocampectomy; ATL = anterior temporal lobe; RNS = responsive neurostimulation; MTS = mesial temporal sclerosis. We reported age ranges to ensure the protection of individual identities.

**Table 2.** PCA Wada test: Wada parameters, effects of drug, Wada memory outcome, and surgical results.

| Patient # | Injection Side/Dose | Hemiplegia | Hemianopia | Aphasia | Memory | Surgery | Engle Score |
| --- | --- | --- | --- | --- | --- | --- | --- |
| 1 | R/60 mg | No | Yes | Normal | Passed | 1) Right SLAH<br>2) Right SAH | 1) 4B<br>2) 1A |
| 2 | BL/50 mg | No | Not Reported | Normal | Passed | L SLAH | 1A |
| 3 | L/60 mg | Mild | Yes | Mild (naming and repetition) | Passed | L SLAH | 1B |
| 4 | L/60 mg | No | Yes | Normal | Passed | L SLAH | 1A |
| 5 | L/50 mg | No | Yes | Normal | Passed | 1) L SLAH<br>2) Repeat | 1) 3A<br>2) 1B |
| 6 | L/60 mg<sup>‡</sup> | No | No | Normal | Passed | L SLAH | 4C |
| 7 | L/50 mg | No | Yes | Normal | Passed | L SLAH | 2D |
| 8 | L/50 mg | No | No | Mild Naming | Passed | L SLAH | 3A |
| 9 | R/60 mg | No | Yes | Normal | Passed | R SLAH | 4A |
| 10 | L/75 mg | No | Yes | Normal | Passed | L SAH | 3A |
| 11 | BL/60 mg | No | Yes | Normal | Passed | L ATL | 1A |
| 12 | L/60 mg | No | No | Normal | Failed | L RNS | 3A |
| 13 | R/50 mg | No | No | Normal | Passed | R RNS | 4A |

Note. <sup>‡</sup>brevital was used instead of sodium amobarbital. PCA = posterior cerebral artery; NP = Neuropsychological; R = Right; BL = Bilateral; L = Left; SAH = selective amygdalohippocampectomy; SLAH = stereotactic laser amygdalohippocampotomy; ATL = anterior temporal lobectomy; RNS = responsive neurostimulation.

### Hypothesis 1: Only a minority of patients who failed the ICA Wada would subsequently fail the PCA Wada

This hypothesis was confirmed. In a group of 13 patients who failed the ICA-Wada, the observed rate of failing the PCA-Wada was 1/13 (8%), significantly less than 50% (*p*=0.003, two-sided binomial test).

### Hypothesis 2. No patients who underwent SLAH after a failed ICA-but passed PCA-Wada would have a catastrophic memory outcome

Clinical lore suggests that patients who fail an ICA-Wada and who have surgery targeting the ipsilateral temporal lobe are at high risk of suffering a catastrophic memory outcome (though precise estimates are difficult to come by). However, in our sample, 0/9 patients who underwent a minimally invasive SLAH after failing an ICA-Wada but then passing a PCA-Wada had a catastrophic postsurgical memory outcome. The observed rate of 0/9 (0%) was significantly less than 50% (*p*=0.004, two-sided binomial test), and our hypothesis was confirmed.

In total, 11/13 patients underwent a destructive procedure (i.e., 9 underwent SLAH and 2 underwent open resections – one ATL and one SAH) with one of these patients experiencing a catastrophic postsurgical decline. This patient (#11 in the tables) experienced a significant decline on verbal memory and language measures following an ATL of the language dominant hemisphere but did not decline on visual memory tasks. As a result, her functional status greatly declined, she lost the ability to live independently, and she has resided in a skilled care facility with memory care for >8 years after undergoing the procedure. She has shown no improvement in memory or language over time. (Tables 3 & 4)

Of the nine patients who failed the ICA-Wada, passed the PCA-Wada, and then underwent SLAH, eight involved the left MTL. The ninth patient underwent right SLAH but had atypical bilateral (left > right) language representation. Of these nine patients, three experienced mild declines in verbal memory (1 SD change), three remained stable, and three showed mild (1 SD change) to large improvements in verbal learning and recall (>2 SD improvements). With regards to visual memory changes in these nine SLAH patients, two experienced moderate to large declines in one of two visual memory tests, while two experienced mild declines in one or both visual memory tests. Four of the nine SLAH patients exhibited mild to large improvement in visual memory following ablation. None of these patients experienced a significant functional decline. Overall, PCA-Wada results were more accurate than the ICA-Wada results for predicting memory outcome after SLAH, as indicated by the mild negative effects on memory tests for most patients, with a significant number of patients experiencing memory improvement after surgery. (Tables 3 & 4)

Of note, the patient with atypical language was not seizure-free following right SLAH, and eventually underwent a right open SAH using a lateral entry approach between the middle and inferior temporal gyri. Though the more extensive procedure yielded seizure-freedom, she then experienced a more significant decline in memory for both auditory/verbal and visual stimuli, which she had not experienced after the SLAH procedure (see patient #1 from Tables 3 and 4). Another patient who failed the ICA-Wada underwent an open left SAH. This patient experienced a large decline in naming and at least a moderate decline in verbal memory, but she did not decline on visual memory measures and was not amnestic.

**Table 3.** Verbal Learning and Memory and Visual Naming outcomes after undergoing TLE surgery following a failed ICA Wada.

| Patient # | Surgery Type | Language Lat | LM I (Pre) | LM I (Post) | LM II (Pre) | LM II (Post) | BNT (Pre) | BNT (Post) |
| --- | --- | --- | --- | --- | --- | --- | --- | --- |
| 1 | Right SLAH | Bilateral (L>R) | 34 (T=47) | 38 (T=50) | 25 (T=50) | 25 (T=50) | 58/60 | 59/60 |
| 1 | Right SAH | Bilateral (L>R) | 38 (T=50) | 38 (T=50) | 25 (T=50) | 28 (T=57) | 59/60 | 58/60 |
| 2 | Left SLAH | Left | 14 (T=30) | 20 (T=40) | 5 (T=27) | 3 (T=27) | 45/60 | 44/60 |
| 3 | Left SLAH | Left | 20 (T=43) | 31 (T=57) | 12 (T=37) | 18 (T=47) | 55/60 | 55/60 |
| 4 | Left SLAH | Left | 23 (T=47) | 22 (T=47) | 26 (T=57) | 15 (T=40) | 37/60 | 39/60 |
| 5 | Left SLAH | Left | 15 (T=30) | 13 (T=30) | 9 (T=26) | 9 (T=26) | 31/60 | 31/60 |
| 6 | Left SLAH | Left | 19 (T=40) | 21 (T=43) | 15 (T=40) | 17 (T=43) | 50/60 | 49/60 |
| 7 | Left SLAH | Left | 24 (T=47) | 20 (T=40) | 17 (T=43) | 11 (T=34) | 45/60 | 53/60 |
| 8 | Left SLAH | Left | 12 (T=27) | 14 (T=30) | 9 (T=30) | 8 (T=27) | N/A | N/A |
| 9 | Right SLAH | Bilateral (L>R) | 19 (T=40) | 21 (T=43) | 17 (T=43) | 16 (T=40) | 42/60 | 47/60 |
| 10 | Left SAH | Left | 26 (T=36) | 5 (T=27) | 7 (T=34) | 0 (T=20) | 38/60 | 29/60 |
| 11 | Left ATL | Left | 15 (T=33) | 5 (T=20) | 8 (T=30) | 5 (T=24) | 41/60 | 18/60 |
| 12 | Left RNS | Left | 20 (T=43) | 17 (T=37) | 16 (T=40) | 14 (T=40) | 31/60 | 30/60 |
| 13 | Right RNS | Right | 37 (T=47) | 21 (T=43) | 26 (T=53) | 10 (T=34) | 47/60 | 42/60 |
| Patient # | Surgery Type | Language Lat | RAVLT 5-Trial (Pre) | RAVLT 5-Trial (Post) | RAVLT Immediate (Pre) | RAVLT Immediate (Post) | RAVLT Delay (Pre) | RAVLT Delay (Post) |
| 1 | Right SLAH | Bilateral (L>R) | 43 (T=41) | 53 (T=52) | 6 (T=33) | 12 (T=56) | 4 (T=28) | 12 (T=56) |
| 1 | Right SAH | Bilateral (L>R) | 53 (T=52) | 36 (T=33) | 12 (T=56) | 10 (T=48) | 12 (T=56) | 9 (T=46) |
| 2 | Left SLAH | Left | 28 (T=26) | 32 (T=31) | 3 (T=38) | 6 (T=51) | 1 (T=26) | 7 (T=41) |
| 3 | Left SLAH | Left | 31 (T=27) | 28 (T=23) | 3 (T=22) | 4 (T=25) | 4 (T=28) | 1 (T=20) |
| 4 | Left SLAH | Left | 56 (T=56) | 58 (T=58) | 9 (T=45) | 6 (T=33) | 10 (T=49) | 6 (T=35) |
| 5 | Left SLAH | Left | 41 (T=31) | 37 (T=24) | 7 (T=32) | 5 (T=17) | 4 (T=16) | 4 (T=16) |
| 6 | Left SLAH | Left | 44 (T=46) | 34 (T=33) | 6 (T=36) | 7 (T=40) | 6 (T=37) | 6 (T=37) |
| 7 | Left SLAH | Left | 35 (T=31) | 28 (T=23) | 5 (T=29) | 4 (T=25) | 5 (T=31) | 2 (T=21) |
| 8 | Left SLAH | Left | 31 (T=23) | N/A | 5 (T=27) | N/A | 5 (T=27) | N/A |
| 9 | Right SLAH | Bilateral (L>R) | 27 (T=18) | 38 (T=35) | 8 (T=38) | 6 (T=33) | 4 (T=25) | 5 (T=31) |
| 10 | Left SAH | Left | N/A | 24 (T=21) | N/A | 0 (T=15) | N/A | 2 (T=25) |
| 11 | Left ATL | Left | 33 (T=32) | 17 (T=12) | 5 (T=36) | 2 (T=26) | 6 (T=38) | 0 (T=18) |
| 12 | Left RNS | Left | 42 (T=36) | 41 (T=35) | 6 (T=31) | 9 (T=45) | 7 (T=35) | 9 (T=46) |
| 13 | Right RNS | Right | 42 (T=36) | 39 (T=34) | 10 (T=46) | 7 (T=36) | 9 (T=43) | 6 (T=35) |

Note. TLE = temporal lobe epilepsy; ICA = internal carotid artery; Language Lat = language lateralization; LM = Logical Memory; BNT = Boston Naming Test; SLAH = stereotactic laser amygdalohippocampotomy; SAH = selective amygdalohippocampectomy; ATL = anterior temporal lobe; RNS = responsive neurostimulation; RAVLT = Rey Auditory Verbal Learning Test; N/A = data not available.
Color coding has been applied to highlight the level of change (both improvement and decline) according to the following key:
**Green** = 1 SD improvement
**Blue** = 2 SD improvement
**Mustard** = 1 SD decline
**Red** = 2 SD decline

**Table 4.**
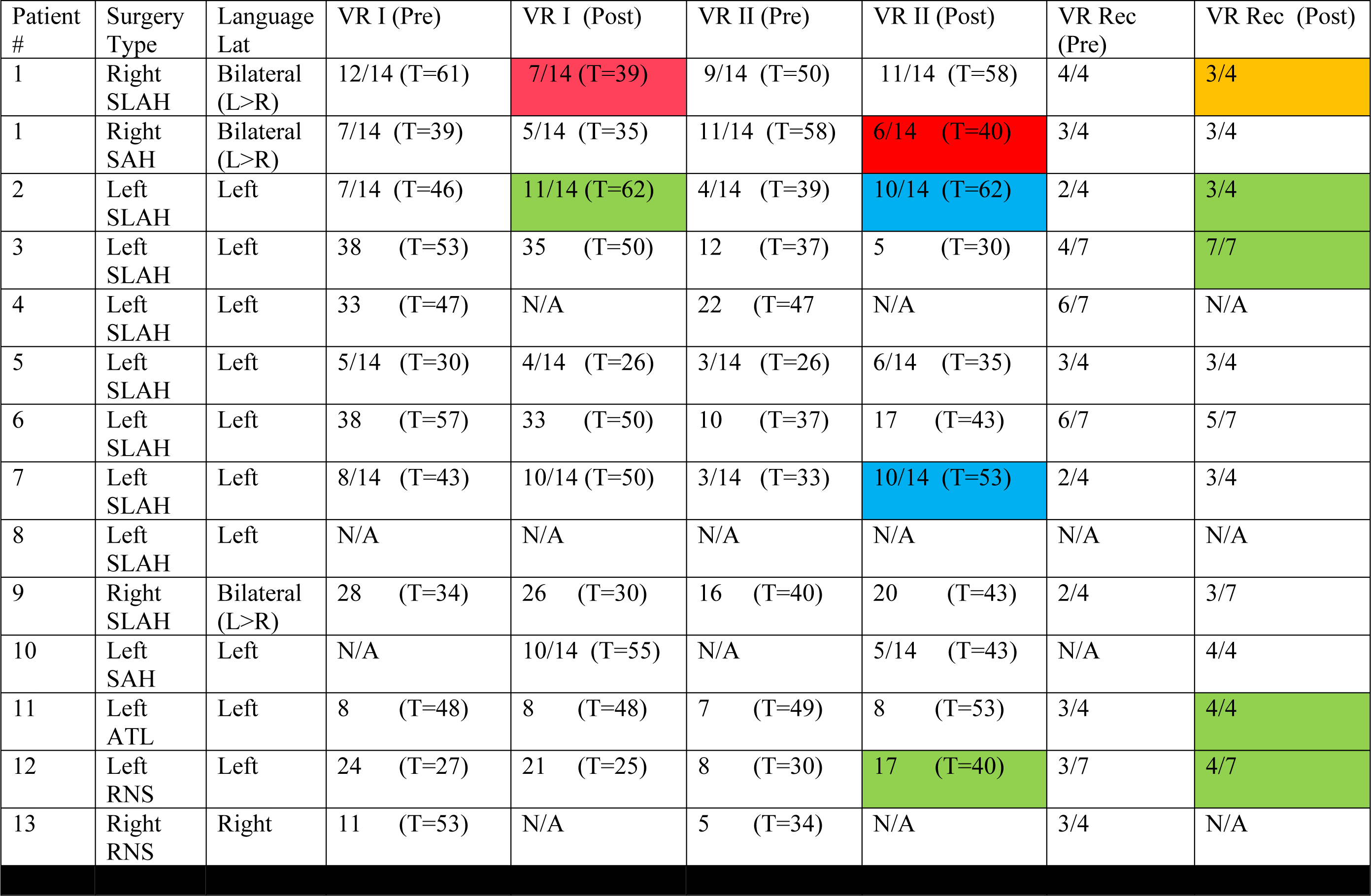
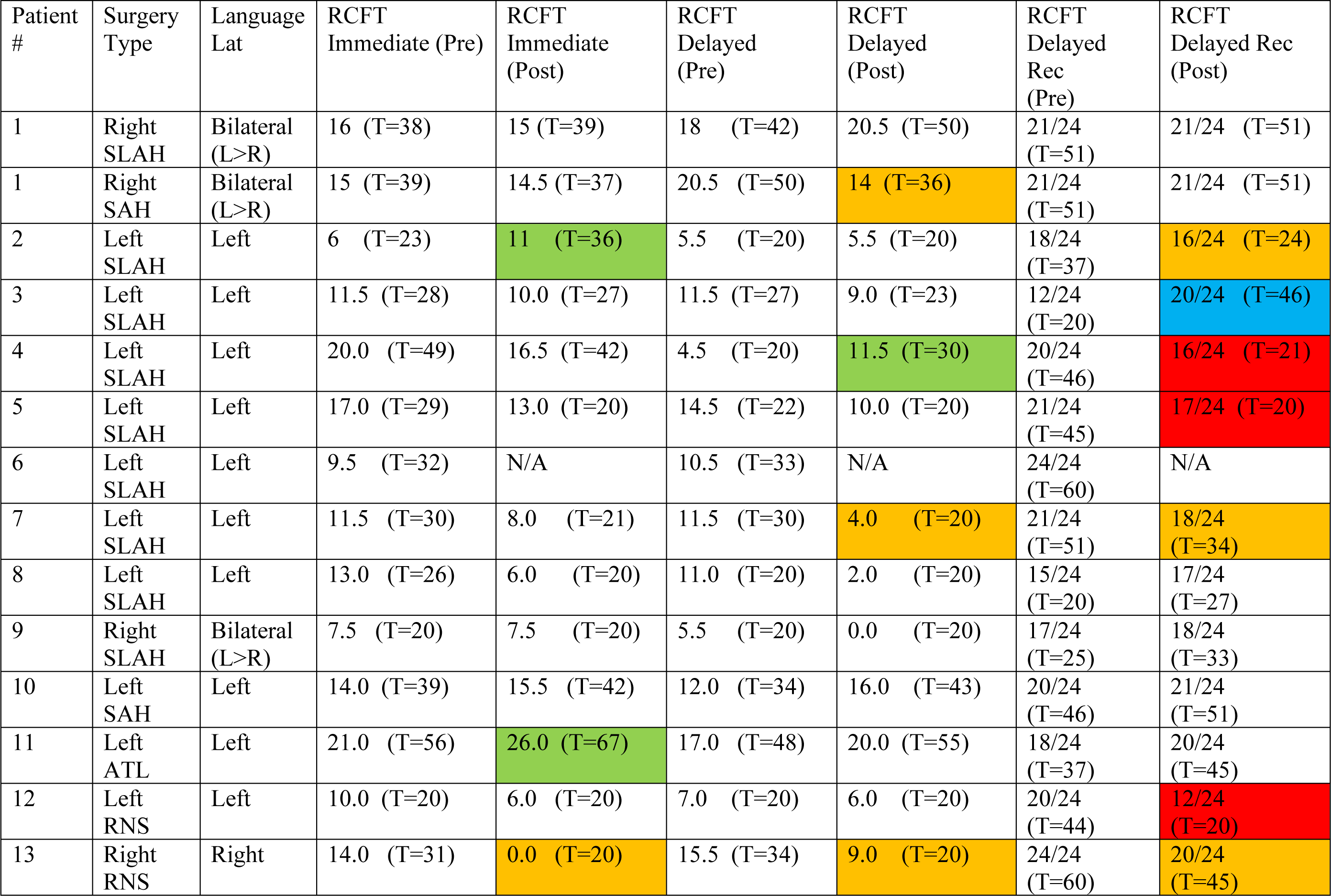
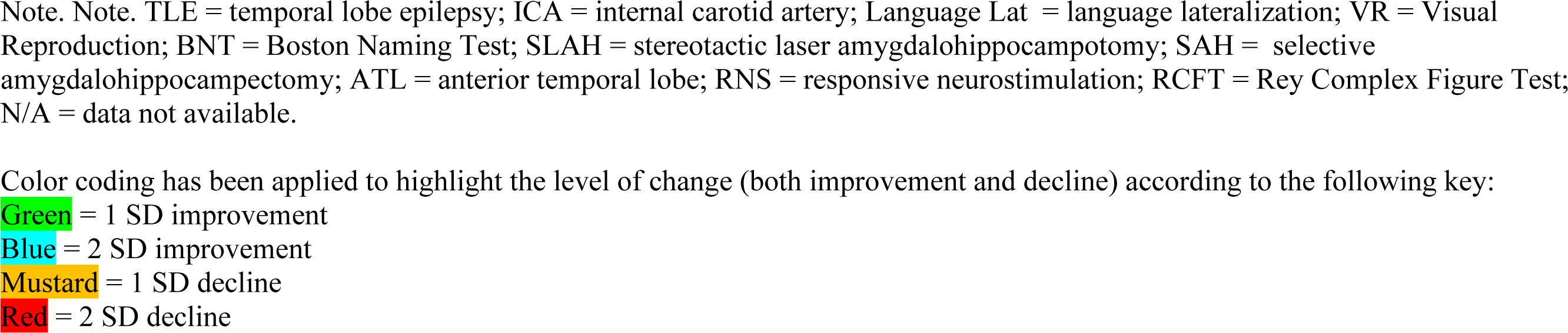
Visual Learning and Memory outcomes after undergoing TLE surgery following a failed ICA Wada.

| Patient # | Surgery Type | Language Lat | VR I (Pre) | VR I (Post) | VR II (Pre) | VR II (Post) | VR Rec (Pre) | VR Rec (Post) |
| --- | --- | --- | --- | --- | --- | --- | --- | --- |
| 1 | Right SLAH | Bilateral (L>R) | 12/14 (T=61) | 7/14 (T=39) | 9/14 (T=50) | 11/14 (T=58) | 4/4 | 3/4 |
| 1 | Right SAH | Bilateral (L>R) | 7/14 (T=39) | 5/14 (T=35) | 11/14 (T=58) | 6/14 (T=40) | 3/4 | 3/4 |
| 2 | Left SLAH | Left | 7/14 (T=46) | 11/14 (T=62) | 4/14 (T=39) | 10/14 (T=62) | 2/4 | 3/4 |
| 3 | Left SLAH | Left | 38 (T=53) | 35 (T=50) | 12 (T=37) | 5 (T=30) | 4/7 | 7/7 |
| 4 | Left SLAH | Left | 33 (T=47) | N/A | 22 (T=47) | N/A | 6/7 | N/A |
| 5 | Left SLAH | Left | 5/14 (T=30) | 4/14 (T=26) | 3/14 (T=26) | 6/14 (T=35) | 3/4 | 3/4 |
| 6 | Left SLAH | Left | 38 (T=57) | 33 (T=50) | 10 (T=37) | 17 (T=43) | 6/7 | 5/7 |
| 7 | Left SLAH | Left | 8/14 (T=43) | 10/14 (T=50) | 3/14 (T=33) | 10/14 (T=53) | 2/4 | 3/4 |
| 8 | Left SLAH | Left | N/A | N/A | N/A | N/A | N/A | N/A |
| 9 | Right SLAH | Bilateral (L>R) | 28 (T=34) | 26 (T=30) | 16 (T=40) | 20 (T=43) | 2/4 | 3/7 |
| 10 | Left SAH | Left | N/A | 10/14 (T=55) | N/A | 5/14 (T=43) | N/A | 4/4 |
| 11 | Left ATL | Left | 8 (T=48) | 8 (T=48) | 7 (T=49) | 8 (T=53) | 3/4 | 4/4 |
| 12 | Left RNS | Left | 24 (T=27) | 21 (T=25) | 8 (T=30) | 17 (T=40) | 3/7 | 4/7 |
| 13 | Right RNS | Right | 11 (T=53) | N/A | 5 (T=34) | N/A | 3/4 | N/A |

| Patient # | Surgery Type | Language Lat | RCFT Immediate (Pre) | RCFT Immediate (Post) | RCFT Delayed (Pre) | RCFT Delayed (Post) | RCFT Delayed Rec (Pre) | RCFT Delayed Rec (Post) |
| --- | --- | --- | --- | --- | --- | --- | --- | --- |
| 1 | Right SLAH | Bilateral (L>R) | 16 (T=38) | 15 (T=39) | 18 (T=42) | 20.5 (T=50) | 21/24 (T=51) | 21/24 (T=51) |
| 1 | Right SAH | Bilateral (L>R) | 15 (T=39) | 14.5 (T=37) | 20.5 (T=50) | 14 (T=36) | 21/24 (T=51) | 21/24 (T=51) |
| 2 | Left SLAH | Left | 6 (T=23) | 11 (T=36) | 5.5 (T=20) | 5.5 (T=20) | 18/24 (T=37) | 16/24 (T=24) |
| 3 | Left SLAH | Left | 11.5 (T=28) | 10.0 (T=27) | 11.5 (T=27) | 9.0 (T=23) | 12/24 (T=20) | 20/24 (T=46) |
| 4 | Left SLAH | Left | 20.0 (T=49) | 16.5 (T=42) | 4.5 (T=20) | 11.5 (T=30) | 20/24 (T=46) | 16/24 (T=21) |
| 5 | Left SLAH | Left | 17.0 (T=29) | 13.0 (T=20) | 14.5 (T=22) | 10.0 (T=20) | 21/24 (T=45) | 17/24 (T=20) |
| 6 | Left SLAH | Left | 9.5 (T=32) | N/A | 10.5 (T=33) | N/A | 24/24 (T=60) | N/A |
| 7 | Left SLAH | Left | 11.5 (T=30) | 8.0 (T=21) | 11.5 (T=30) | 4.0 (T=20) | 21/24 (T=51) | 18/24 (T=34) |
| 8 | Left SLAH | Left | 13.0 (T=26) | 6.0 (T=20) | 11.0 (T=20) | 2.0 (T=20) | 15/24 (T=20) | 17/24 (T=27) |
| 9 | Right SLAH | Bilateral (L>R) | 7.5 (T=20) | 7.5 (T=20) | 5.5 (T=20) | 0.0 (T=20) | 17/24 (T=25) | 18/24 (T=33) |
| 10 | Left SAH | Left | 14.0 (T=39) | 15.5 (T=42) | 12.0 (T=34) | 16.0 (T=43) | 20/24 (T=46) | 21/24 (T=51) |
| 11 | Left ATL | Left | 21.0 (T=56) | 26.0 (T=67) | 17.0 (T=48) | 20.0 (T=55) | 18/24 (T=37) | 20/24 (T=45) |
| 12 | Left RNS | Left | 10.0 (T=20) | 6.0 (T=20) | 7.0 (T=20) | 6.0 (T=20) | 20/24 (T=44) | 12/24 (T=20) |
| 13 | Right RNS | Right | 14.0 (T=31) | 0.0 (T=20) | 15.5 (T=34) | 9.0 (T=20) | 24/24 (T=60) | 20/24 (T=45) |

Note. Note. TLE = temporal lobe epilepsy; ICA = internal carotid artery; Language Lat = language lateralization; VR = Visual Reproduction; BNT = Boston Naming Test; SLAH = stereotactic laser amygdalohippocampotomy; SAH = selective amygdalohippocampectomy; ATL = anterior temporal lobe; RNS = responsive neurostimulation; RCFT = Rey Complex Figure Test; N/A = data not available.
Color coding has been applied to highlight the level of change (both improvement and decline) according to the following key:
**Green** = 1 SD improvement
**Blue** = 2 SD improvement
**Mustard** = 1 SD decline
**Red** = 2 SD decline

The single subject (patient #12) who failed both the ICA- and PCA-Wada was offered a neuromodulatory intervention (i.e., unilateral left responsive neurostimulator [RNS] implantation with hippocampal depth electrodes, rather than a destructive procedure) and remained cognitively stable postoperatively. Because we sought to avoid catastrophic memory outcomes and did not offer a destructive procedure after failed PCA-Wada, we cannot examine the positive predictive value of a failed PCA-Wada upon memory outcome.

There was one additional case of unilateral RNS implantation (patient #13) after failed ICA-Wada. In this case, the patient selected RNS over SLAH due to fear of a negative cognitive outcome, despite having passed the PCA-Wada. Notably after RNS placement with two hippocampal electrodes, the patient experienced mild to large declines on more than one memory measure, which appeared to be driven by declines in general processing speed, attention (severe deficits), and level of alertness. She eventually had the unilateral RNS device removed, and her cognitive functions returned to preimplantation baseline.

### Hypothesis 3: Language disturbance will not be seen in patients who undergo SLAH but will instead be observed following open resection procedures that extend beyond the MTL

No SLAH patients declined on naming ability following surgery (0/9), while two of three open resection patients experienced significant decline (2/3) (Fisher’s exact test, *p*=0.046). Additionally, two of nine SLAH patients experienced significant improvement on the visual naming test following surgery.

We compared the percent decline pre- and post-surgery in raw scores in several neuropsychological tests, comparing SLAH versus open surgery. There was a statistically significant difference between the percent decline for logical memory: Logical Memory I (LMI, +12% SLAH vs. -49% Open, *p*=0.009, unpaired t-test), Boston Naming Test (BNT, +4% vs. -27%, *p*=0.01, unpaired t-test), and Rey Auditory Verbal Learning Test 5-trial (RAVLT 5, +2% vs. -40%, *p*=0.03, unpaired t-test). There were no significant differences between the percent decline for SLAH vs. Open for the remaining verbal tests (Figure 3 top), or any of the 6 visual tasks (Figure 3 bottom).

**Figure 3.**
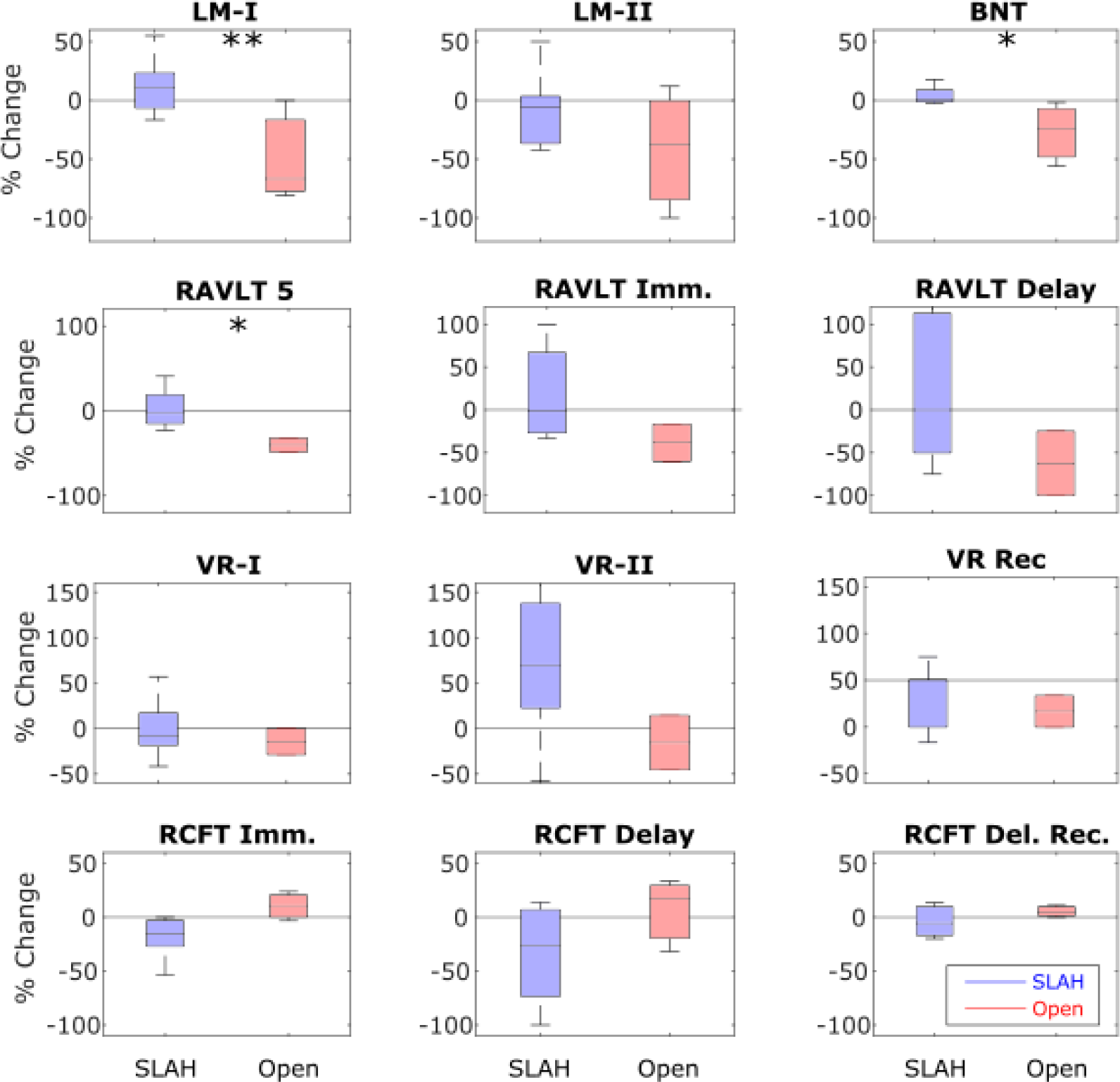
Percent change in neuropsychological tests before and after SLAH versus open TL surgical procedures. Box plots are shown for the % change in the raw scores of individual tests. An unpaired t-test was used to compare SLAH vs. open procedures (* is p<0.05, ** is p<0.01). Abbreviations are listed in the legend of Table 2 and 3.

**Figure 4.**
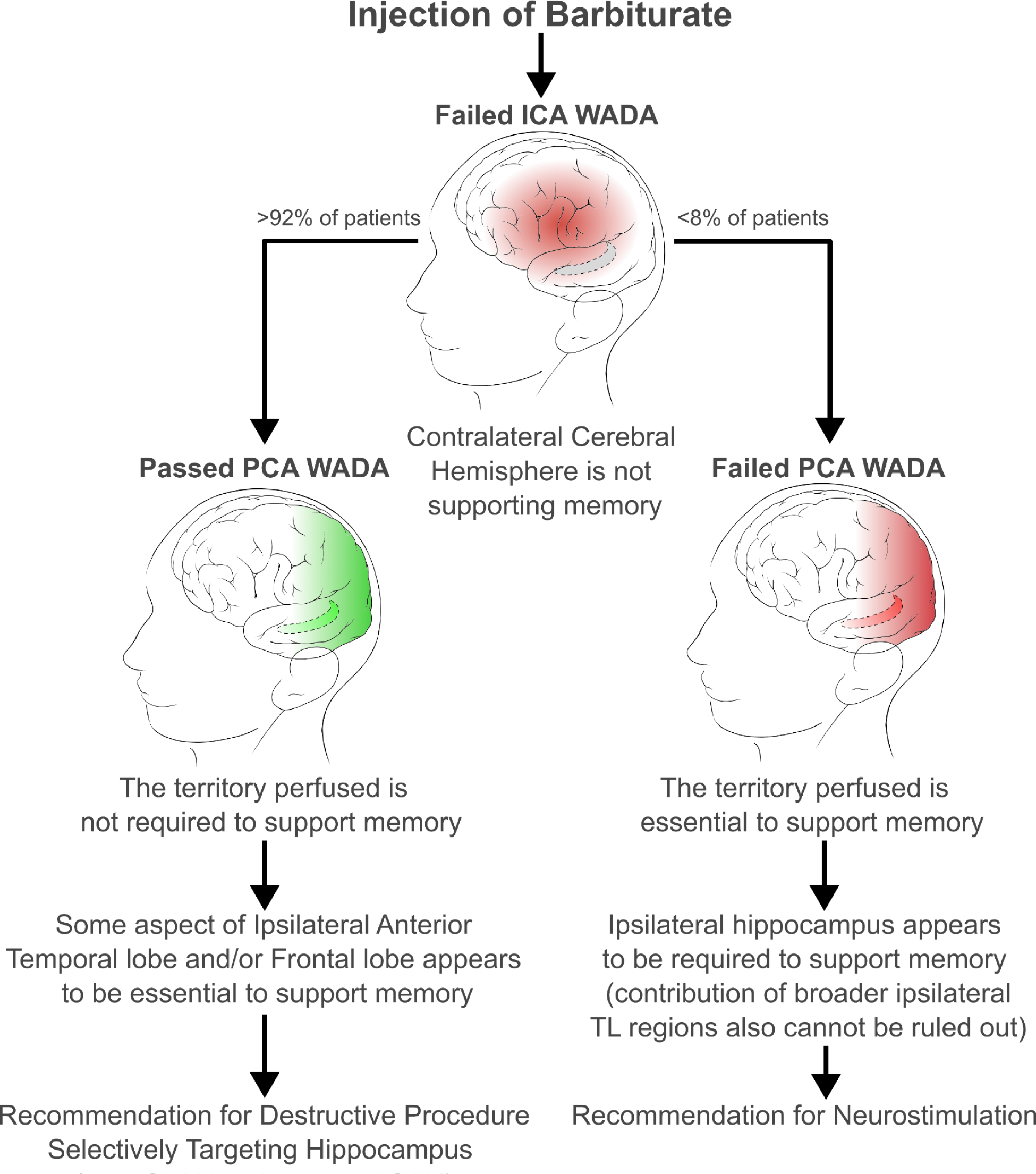
Surgical recommendations based upon ICA- and PCA-Wada results. When a patient “fails” the ICA Wada for memory (i.e., the patient fails to subsequently recognize objects presented during brain inhibition by barbiturate), after a recovery interval, a PCA injection is administered, and an analogous recognition memory test performed to evaluate the contribution of the ipsilateral hippocampus to memory. In >90% of cases of failed ICA-Wada, the patient subsequently passed the PCA-Wada, suggesting that the ipsilateral hippocampus was not essential to memory. Thus, such patients can undergo highly selective approaches to the MTL, such as SLAH. For the patients that failed the PCA-Wada, this is interpreted to mean that the ipsilateral hippocampus is required for recognition memory, and thus neuromodulation is generally recommended as first line therapy over destructive procedure.

## Discussion

Our most striking observation was that no MTLE patients who passed a PCA-Wada test for declarative object memory (despite having failed the ICA-Wada test for memory) exhibited catastrophic memory outcomes following subsequent SLAH. By contrast, a patient who underwent language dominant hemisphere ATL after having failed both PCA- and ICA-Wada tests had a near catastrophic memory outcome. Likewise, while ICA-Wada is a gold-standard for determining the hemisphere of language dominance, ipsilateral PCA-Wada rarely impacts language. Indeed, the patients who underwent SLAH after PCA-Wada tests exhibited no significant decline in confrontational naming. Our findings suggest that while the ICA-Wada may help predict risk of memory and naming declines after open ATL, it yields a false positive risk detection for such deficits following SLAH. Thus, passing the PCA-Wada of declarative object memory better predicts memory outcomes in patients undergoing SLAH.

Overall, the concordance of results between PCA-Wada and SLAH are consistent with the notion that in MTLE, the amygdalo-hippocampal complex can be treated without particular risk of catastrophic memory decline if the ipsilateral temporal lobe neocortex and contralateral structures remain functionally intact. Likewise, the discordance of results of ICA-Wada to those of SLAH indicate that avoiding collateral injury to ipsilateral neocortex is likely critical to maintaining memory and naming functions. These observations highlight the need for personalized pathological and functional mapping of seizure networks and cognitive dependencies, especially when considering destructive procedures with variable impact upon off-target structures. The importance of anterior-lateral and polar TL neocortices outside the MTL to memory and learning remain insufficiently considered in the clinical setting. While the ICA-Wada may predict deficits after open ATL, subsequent passing the PCA-Wada test for declarative object memory may allow patients to still undergo SLAH even when considered to be at high risk from open ATL.

Nearly all patients who failed the ICA-Wada went on to pass the PCA-Wada for declarative object memory. The single patient who failed both Wada procedures was treated with a neuromodulation device. Because open ATL and SLAH may be contraindicated in patients who fail the ICA- and PCA-Wada tests, respectively, determining the positive and negative predictive values for each tests with respect to each surgical approach is needed, but likely impractical. Our results do suggest that the false-positive outcome risk of the conventional ICA-Wada test is high for the SLAH procedure, because the brain regions anesthetized are broader that tissues targeted by SLAH (i.e. diagnostic-therapeutic mismatch). Overall, patients in whom only the MTL are to be targeted should not be disqualified from SLAH based solely upon ICA-Wada results. On the other hand, because the ICA distribution does not reach much of the hippocampus targeted by ATL and SLAH,^11,12^ it is also possible that the ICA-Wada could produce false-negative outcome risks for these standard surgical approaches. The PCA-Wada fills this gap and may be a more precise approach to assessing cognitive risk of removing the MTL, especially the hippocampal complex, prior to surgical procedures (i.e. good diagnostic-therapeutic match).

We observed that the ICA- and PCA-Wada tests produce vastly different results with respect to declarative memory for objects, and that the deficits generally induced by the ICA-Wada test are inconsistent with the neurocognitive outcome of the SLAH procedure. These findings concord well with the only other direct comparison of ICA- and PCA-Wada results, which were obtained in the context of evaluation of candidates for open subtemporal SAH procedures at another center.^29^ That group similarly demonstrated an absence of adverse memory change in 13 patients who underwent a subtemporal SAH despite a failed ICA-Wada.

Nevertheless, we caution that due to natural variations in individual vascular anatomy (see supplemental figure 1), behavioral results should be interpreted in the context of individual angiographic results. While PCA branches likely perfuse more of the the hippocampus in most individuals, there are patients for whom ICA branches perfuse predominantly. Indeed, the variable blood supply of the hippocampal head is consistent with it being within a vascular watershed area that is sensitive to ischemic injury.

The patient in our study who failed the ICA Wada but passed the PCA Wada and went on to receive a left language dominant ATL experienced a near catastrophic memory outcome, which hints at the positive predictive value for the ICA Wada in the setting of ATL. Following surgery, this patient never returned to work, went through a lengthy period of neurologic rehabilitation, and experienced significant depression. She required assistance with most life tasks, and rapidly entered an assisted living facility with memory care. Now more than eight years after her surgery, her mood has stabilized. Still, she remains dependent on others for her care and continues to exhibit burdens created by poor memory and naming (e.g., she has been unable to learn the names of her grandchildren or new persons she meets, cannot manage her finances, and needs help with medications). Notably, she was not fully amnestic, as non-verbal memory did not decline, and she still retained some gist of her larger circumstances, suggesting that she still had some limited capacity for autobiographical memory. Primarily, she has experienced significant impairment of verbal learning and memory and language skills. Overall, these findings highlight that the ICA Wada demonstrates the effects of anesthetizing a large extent of the anterior TL region rather than its isolated effects on the hippocampus. These findings also suggest that it was the extra-hippocampal networks of this patient’s brain that were sustaining critical verbal memory and language function. Despite being counseled about the high risk of an unfavorable outcome in terms of language and memory (including a possible risk of becoming amnestic), the patient requested the left ATL procedure to maximize the chances of seizure relief in a single surgery, given concerns about the ongoing effect of seizures on her daily life and overall health. Additionally, she made this choice during the first year of our experience with SLAH, and cognitive outcomes for this procedure were still unknown to us.

While the focus of this study is upon predicting direct cognitive outcomes of temporal lobe surgery, we recognize that different surgical approaches yield different rates of seizure freedom, which will also indirectly impact cognitive outcomes. In cases in which minimally invasive SLAH does not achieve seizure freedom, there are instances in which a subsequent wider resection or ablation, despite additional cognitive risk, is necessary to achieve optimal seizure control. Given the reported results in subtemporal SAH procedures with regard to potentially preserving memory in a prior study,^29^ we recommend a case-controlled direct comparison of seizure and cognitive outcomes between these two distinct approaches. Published outcome data regarding the subtemporal SAH are generally limited, and there are no studies directly comparing subtemporal SAH to SLAH or any of the other open selective approaches. We note that our study identified significant memory decline in two cases following an open SAH approach that transected the lateral temporal lobe via the inferior temporal sulcus. This further highlights that differential outcome patterns are likely even across variant approaches to SAH.

We emphasize that several patients in our study who were initially considered at risk of severe postsurgical memory deficits based on the ICA-Wada test actually showed significant memory improvement following minimally invasive SLAH, or in fewer cases, following open selective resection. It is reasonable to assume that the memory improvement in these patients resulted from the virtuous combination of seizure control and sparing of cognitively important lateral and anterior temporal structures, maximizing patient capacity for cognitive recovery.

This work has some limitations. Larger sample sizes, comparison of patient characteristics to individual memory outcomes, and a better understanding of variability of amobarbital distribution following PCA injection are needed to further characterize the utility of this approach. In addition, other fundamental variables in relation to outcome need to be explored: (1) are Wada results skewed by poor memory at baseline? And (2) are failed Wada results more meaningful when patients have strong evidence of bitemporal disease (e.g., structural abnormalities, bilateral PET hypometabolism, or causes of epilepsy from encephalitis or TBI with higher risk of bilateral injury)? As described above, we were particularly unable to evaluate the positive predictive value of a failed PCA-Wada, given only one participant failed and did not undergo destructive surgery.

Despite fears about the technical safety of the PCA-Wada by some in the field,^37,38^ our study demonstrates that this procedure can be performed regularly without adverse complications. The risk of the PCA-Wada appears to be low when implemented by experienced angiographers. With regard to the study effects of the PCA-Wada, we note that it rarely resulted in the hemiplegia or aphasia typical of the ICA-Wada. This is important for both clinical and neuroscientific reasons. From a clinical perspective, the PCA-Wada is inadequate to assess language or motor function, and is thus more dependent upon concurrent EEG to verify anesthetic effects on the brain. In contrast, the ICA-Wada will lead to deficits in both motor and language functions, again highlighting its broad effect upon the perisylvian frontal and temporal regions. From a neuroscience perspective, our data support mounting evidence that the hippocampus is not significantly involved in language functions that are tested in Wada’s procedure (at least in the context of medial temporal lobe epilepsy). In the research literature, many have proffered a role for the hippocampus in the retrieval of words and names as well as other language functions, usually on the basis of correlated electrophysiological or structural neuroimaging findings.^39-41^ However, our data clearly demonstrate an absence of most language findings during hippocampal anesthetization, and we observed no language deficits after any of the nine SLAH procedures performed in this cohort, but naming declined severely in the two patients who underwent larger open resections. These results concord with behavioral outcomes we have described previously (i.e., naming decline does not occur following SLAH despite frequent occurrence following open ATL and/or SAH).^25,42^

We have recently described efforts to utilize direct electrical stimulation (DES) to directly disrupt hippocampal function and evaluate memory–an “electric Wada.”^30^ A subject who underwent evaluation for likely left TLE failed a conventional ICA-Wada but was then unable to tolerate a subsequent PCA-Wada. She underwent intracranial stereoelectroencphalography (SEEG) which localized seizures to the left hippocampus, and we then used a variety of stimulation parameters across multiple hippocampal contacts to seek to disrupt memory function in the absence of eliciting seizures. When we found no acute change in memory function with stimulation mapping, the hippocampus was ablated in two stages over time, initially with a partial radiofrequency thermal ablation via the SEEG depth electrodes contacts. After an interval of some months without detection of memory or other cognitive decline and when seizures recurred, we carried out a more definitive SLAH. This patient has subsequently been seizure-free for nearly two years and showed no negative declines in performance. Additional work will be required to determine if direct electrical stimulation protocols can be developed to reliably test the memory functions of target tissue. Ultimately there is a need to establish methods of acutely disrupting memory with one or more of these methods (PCA-Wada, electric Wada, etc.), and determine which structures/regions must be affected to cause memory change in varying clinical contexts. It will also be important to extend the array of cognitive, language, sensory processing, and socio-emotional tasks to better determine causality (necessity and sufficiency) of particular structure-function relationships within the temporal lobe.^43^

An emerging noninvasive brain mapping method utilizes temporally interfering electrical fields (TIEF),^44-46^ which may affect very focal regions of brain tissue, such as hippocampus, in an effort to simulate the effects of surgery across cognitive or socio-emotional tasks. A more flexible method of noninvasive stimulation would potentially supplement or replace DES mapping, such as for the assessment of language,^23^ and extend the reach of noninvasive mapping to subcortical structures. DES techniques currently rely on invasive methods often constrained by the typically sparse placement of SEEG electrodes. Other non-invasive stimulation tools, such as transcranial magnetic (TMS) or alternating current stimulation (tACS) have a limited spatial ‘reach’ and do not target deep cortical and subcortical structures with spatial specificity.^47-50^ Indeed, insufficient focality relative to DES has been a source for concern.^50^ TMS and tACS cannot be used to focally target the hippocampal formation. In contrast, TIEF holds the promise of being able to reach deep brain structures, and it appears that the delivery envelope of electricity can be modified to achieve both focal precision or broader disruption as required by the goal of clinical or research inquiry. We foresee eventual replacement of conventional pharmacological Wada tests with less invasive and more precise mapping techniques. Overall, the future of determining precise risk-benefit calculations for each possible surgical option and situation will depend upon developing the optimal tools to establish detailed structure- or network-function relationships.

## Data Availability

All data produced in the present study are available upon reasonable request to the authors.

## Acknowledgements

This work was in part supported by grant funding (R01 NS088748, K02 NS0709060) received by Dr. Daniel L. Drane from the National Institute of Neurological Disorders and Stroke (NINDS) of the National Institutes of Health (NIH). NPP was supported by CURE Epilepsy, and NINDS K08NS105929, and R21NS122011. Jon T. Willie is supported by R01MH120194 (NIMH) and P41EB018783 (NIBIB).

## Footnotes

1 We should note that the use of the ICA and PCA Wada procedures was always determined on clinical grounds in an effort to calculate the risk-benefit of the proposed surgical procedure. Our retrospective imposition of hypotheses on our dataset was done to allow us to formulate a statistical test of our outcomes, and should not be taken as the treatment teams’ approach to the individual patient.

## Supplemental Materials

The blood supply to the medial temporal lobe and hippocampus can be highly variable with contributions from branches of the anterior choroidal artery from the ICA and the hippocampal artery or arteries, which may derive from the cisternal segment of the PA directly, or as a single or multiple branch of any of the inferior temporal arteries. The latter include the anterior, middle, and posterior temporal arteries. Therefore, our findings may vary by patient, and individual anatomy can be an important factor to consider at the individual patient level. For this manuscript, we are trying to make the general point that brain perfusion from the ICA injection is typically much broader than than from the PCA injection.

**Figure S1.**
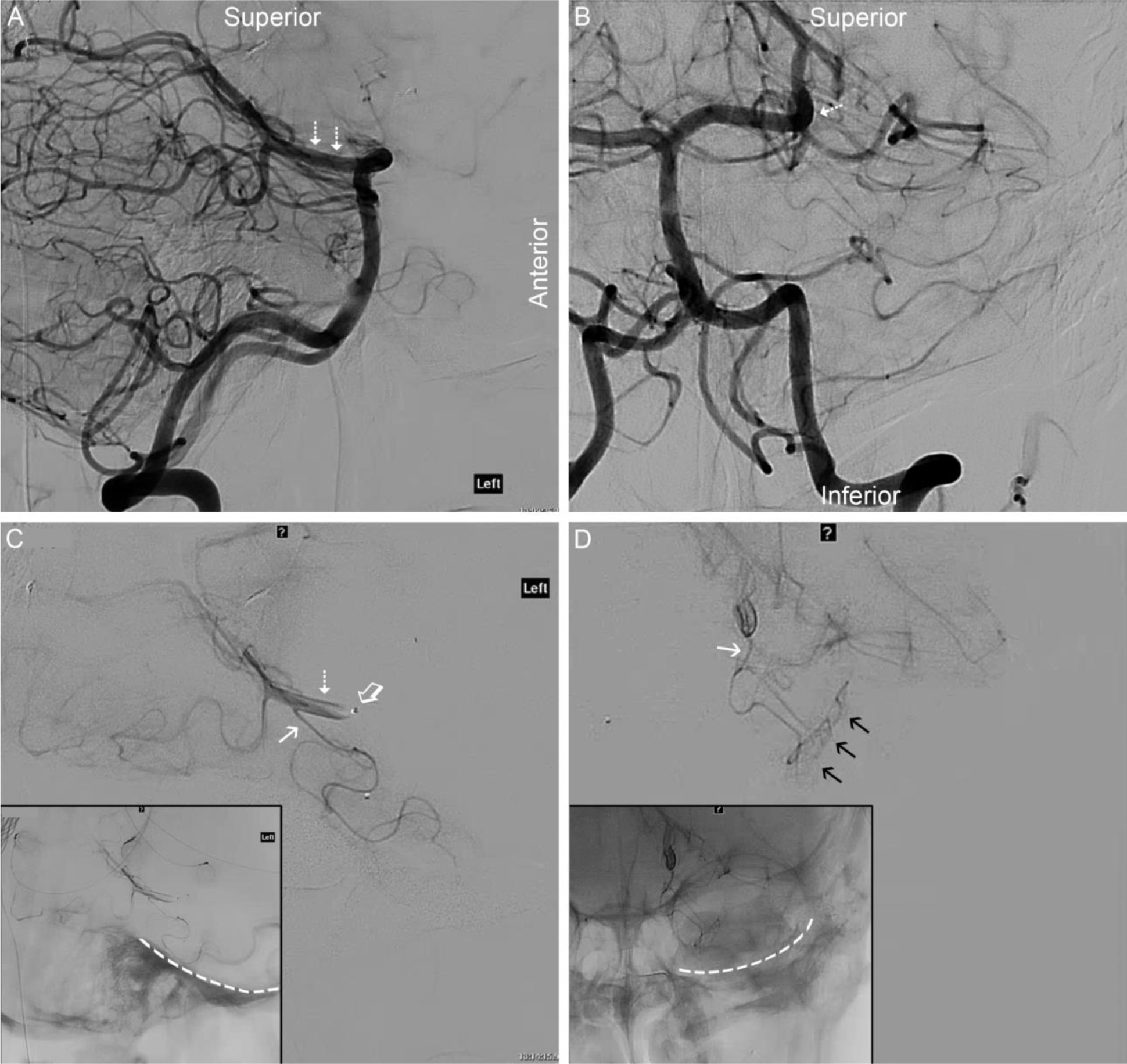
Angiographic Anatomy of the PCA WadaTest. A) Lateral angiogram of the anterior circulation via an ICA injection. The anterior choroidal artery (AChorA) is, typically, the distal-most branch of the supraclinoid ICA (open black arrow). As AChorA enters the ventricular system at the anterior choroid plexal point (jagged white arrow), it makes a sharp turn before straightening in its course along the choroid plexus of the temporal horn of the lateral ventricle (black dashed arrow). B) anterior-posterior (AP) angiogram of the anterior circulation. The AChorA and medial temporal lobe blush are difficult to distinguish on ICA injections. C) Lateral angiogram of the posterior circulation. Dashed arrows denote the P2 or cisternal segment of the PCA. D) Focused AP angiogram of the posterior circulation. The dashed arrow again denotes the mid-P2 segment. E) Selective, lateral angiogram from the microcatheter, which is situated in the mid P2 segment, proving that the microcatheter (tip marked by the open arrow) is distal to brainstem and thalamic perforators. F) Selective, AP angiogram from the mid-P2 segment. In this patient, a single, dominant inferior temporal artery is noted (solid arrow), off which the hippocampal artery arises. In the AP view, the hippocampal artery is seen giving rise to the typical hippocampal arcade immediately medial to the temporal horn of the lateral ventricle (black arrows). E, F Insets) Unsubtracted angiograms reveal the relevant bony anatomy of the middle fossa. The dashed, white line depicts the bony floor of the middle fossa.

